# Childhood predictors of dementia and mortality: 78 year follow-up of the 1946 British Birth Cohort

**DOI:** 10.64898/2026.03.03.26347490

**Authors:** Josh King-Robson, Jennifer M Nicholas, Sarah-Naomi James, James W Groves, Nish Chaturvedi, Marcus Richards, Jonathan M Schott

## Abstract

**Background:** Early-life origins of late-life health are unclear. It is uncertain when in life risk-factors for dementia begin to exert effects. We examined childhood predictors of dementia and mortality in the world’s longest continuously-running birth cohort.

**Methods:** Prospective cohort study, 78-year follow-up. Exposures assessed from birth-age 16. Dementia/death identified from national registries. Cox regression identified childhood predictors.

**Findings:** Of 5,046 participants, 121 (2·4%) were diagnosed with dementia and 1,243 (24·6%) died by age 78.

Independent childhood predictors of dementia were low cognition (HR 2·06, 95% CI 1·31-3·24, *P*=0·002), pollution exposure (HR 2·05, 1·15-3·65, *P*=0·02), left-handedness/ambidexterity (HR 1·93, 1·15-3·23, *P*=0·01), sleep disturbance (HR 1·78, 1·00-3·17, *P*=0·049), nocturnal enuresis (HR 1·52, 1·06-2·17, *P*=0·02), and emotional problems (HR 1·38, 1·12-1·69, *P*=0·003). *APOE* L4 (HR 3·38, 1·67-6·85, *P*=0·001), and BMI (HR 1·47, 1·17-1·84, *P*<0·001) associated with dementia only in females.

Independent childhood predictors of mortality were BMI (HR 1·07, 1·01-1·14, *P*=0·02), lower socioeconomic position (HR 1·25, 1·09-1·42, *P*=0·001), sleep disturbance (HR 1·27, 1·05-1·53, *P*=0·02), nocturnal enuresis (HR 1·32, 1·04-1·68, *P*=0·02) emotional problems (HR males:1·09, 1·00-1·18, *P*=0·046, females:1·25, 1·14-1·36, *P*<0·0001), and conduct problems (HR 1·13, 1·06-1·20, *P*=0·0002). Female sex (HR 0·67, 0·59-0·75, *P*<0·001), education (HR 0·76, 0·67 to 0·87, *P*<0·001), and breastfeeding (HR 0·93/3-months, 0·89-0·98, *P*=0·002) associated with lower mortality.

**Interpretation:** Childhood factors predict a significant proportion of dementia and early mortality risk. While some associations are genetically determined or may relate to early brain development, the majority are potentially modifiable.

**Funding:** Medical Research Council, Alzheimer’s Research UK, Alzheimer’s Association, NIHR, UK Dementia Research Institute.

## Introduction

The early years of life are pivotal to human health and development, yet little is known about the longer-term consequences of childhood characteristics, exposures, and attainment on late-life brain health and mortality. Dementia is the leading cause of death in the UK.^1^ Of fourteen potentially modifiable risk factors for dementia proposed by the Lancet Commission, education was deemed the only early-life factor strongly associated with dementia risk.^2^ While mid– or late-life exposure to other modifiable risk factors including obesity, impaired sleep, mood, vision, and hearing are associated with dementia and mortality,^2–6^ lack of long-term data in cohorts studied since birth means that it is unclear if these associations exert influence even earlier in life. Furthermore, the relatively short follow-up of most studies and co-association with other health predictors often complicates their interpretation. Particularly when considering dementia risk, it is unclear whether risk factors identified even many years prior to diagnosis are manifestations of prodromal disease or contribute to neurodegenerative processes.^2,7^

The concepts of cognitive reserve and resilience provide an important framework to understand differences in susceptibility to dementia risk, and capture the capacity of the brain to maintain cognition and function in the face of ageing and disease. Within this framework, brain reserve reflects structural capacity, largely shaped during early development; brain maintenance refers to modifiable changes that affect brain structure and pathology over time; and cognitive reserve reflects the efficiency and flexibility of the brain to preserve cognitive performance despite brain insults, including the pathologies that cause dementia.^2,8^ Cognitive reserve may be influenced by education, but other exposures associated with improved health and cognition likely also exert positive influence. Conversely, subtle developmental differences and negative health exposures may increase risk both for dementia and premature mortality.^2,8–10^

Identifying associations between childhood factors and late-life health outcomes has major implications for understanding causal pathways and designing public health interventions. The prospective evaluation of these relationships is precluded by need for a lifetime of follow-up. In this study we harnessed data from the world’s longest continuously-studied birth cohort to examine the relationship between childhood factors, dementia risk, and mortality over 78 years follow-up.

## Methods

The MRC National Survey of Health and Development (NSHD) initially comprised 5,362 participants born in mainland Britain in the same week in March 1946.^11^

### Protocol approvals, registrations, and patient consents

Required approvals obtained, most recently: National Research Ethics Service Committee London (REC reference:14/LO/1173). All participants provide written informed consent.

### Childhood predictors

Predictors were selected, where data were available from childhood, to include all potential (sleep) or established modifiable risk factors for dementia (less education, hearing impairment, depression, obesity, air pollution, visual impairment) identified by the Lancet commission at any point in the life course.^2^ We additionally selected non-modifiable risk factors with established associations with dementia risk including sex, Apolipoprotein E *(APOE)* L4 carrier status, socioeconomic position, cognition.^2,12^ Developmental delay, handedness and nocturnal enuresis were included as potential markers of altered early brain development, alongside birthweight and breastfeeding due to their strong associations with cognitive and general health outcomes.^2,9,10,13^ Detailed in **appendix [p2-3]**.

### Mortality and dementia

Mortality data and date of emigration for England and Wales were collected from the NHS central register to age 78·3 years. Date at first dementia diagnosis was determined from nationally collected Hospital Episode Statistics (HES) to age 78·0 (**appendix [p3]**).

### Statistical analysis

All participants with diagnostic and mortality data were included in analysis.

Cox proportional hazards regression examined the relationship between childhood factors and risk of mortality and dementia diagnosis by age 78 (**appendix [p3]**). All analyses were adjusted for sex. We additionally examined for an interaction between sex and each childhood factor in their relationship with dementia and mortality. *P*<0·05 was considered statistically significant.

#### Multivariable analyses

Factors showing a relationship with mortality or dementia diagnosis (*P*<0·1) were selected for inclusion in multivariable Cox proportional cause-specific hazards models, including significant interactions with sex. Backwards stepwise elimination was then used to exclude predictors which no longer remained statistically significant (*P*<0·05). To mitigate potential bias from missing data all multivariable analyses used multiply imputed datasets. An exploratory post-hoc analysis calculated the population attributable fraction (PAF) for mortality and dementia diagnosis by age 78 (**appendix [p3-6]**).

Adjusted survival plots were predicted from regression models (**appendix [p6]**).

Sensitivity analyses were performed for confirm linearity assumptions, identify whether mortality results were driven by childhood mortality, significant associations with BMI by age at menarche, or nocturnal enuresis by those with persistent nocturnal enuresis throughout childhood, and examined accuracy of handedness assessment (**appendix [p7]**).

## Results

### Participant characteristics

5,046 NSHD participants had diagnostic and mortality data available; 121 (2·4%) were diagnosed with dementia and 1243 (24·6%) died by age 78. Participant characteristics are summarised in **Table 1**. Data availability for each exposure is listed in **Figures 1 and 2**.

**Figure 1:**
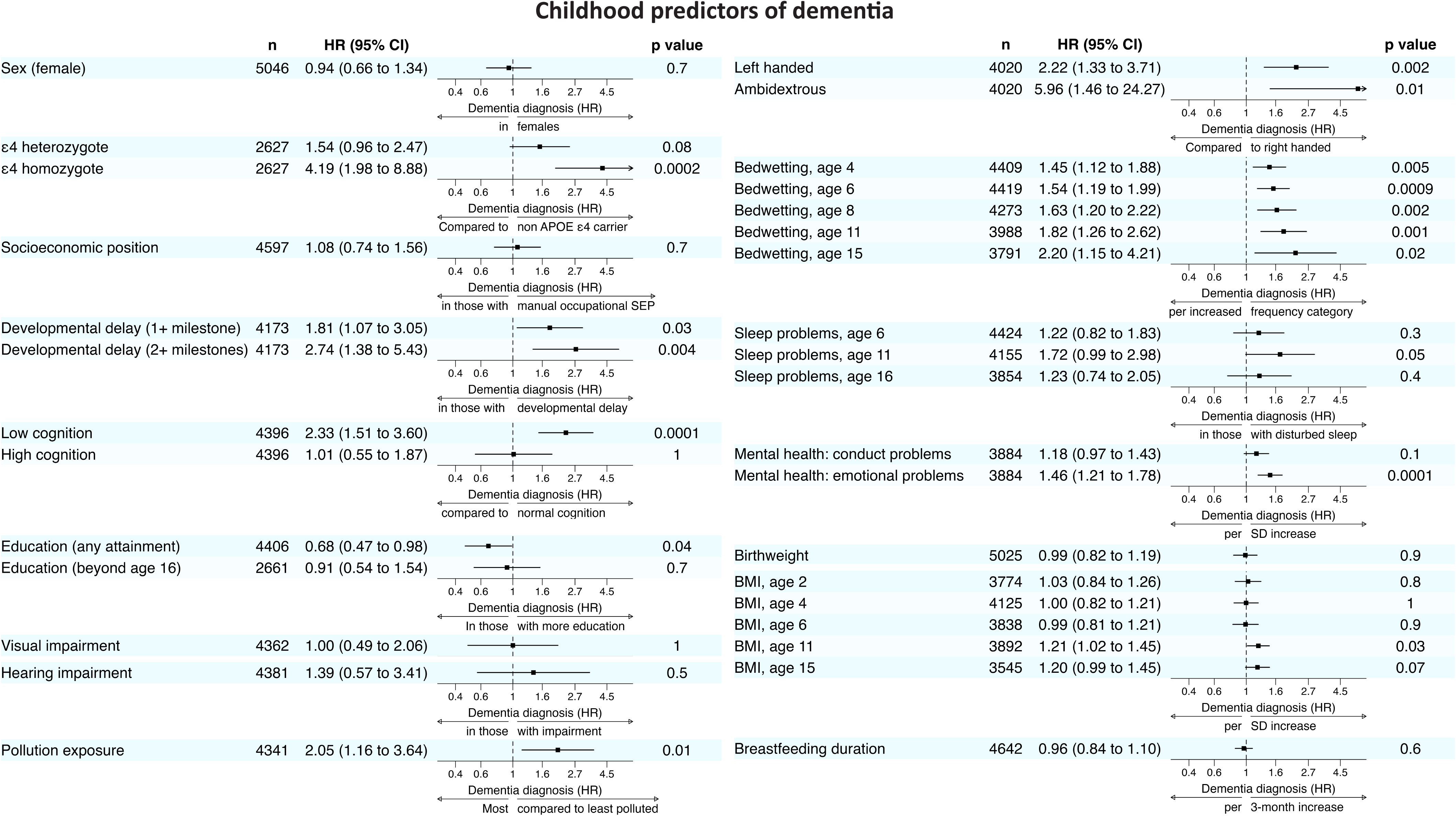
Childhood predictors of dementia. Forest plots demonstrating results of Cox proportional cause-specific hazards regression to examine the relationship between childhood factors and dementia diagnosis by age 78, censoring for emigration and the competing risk of death, and adjusting for sex. Education beyond age 16 is compared only to those with any educational attainment up to age 16. HR; hazard ratio (per unit change, for continuous variables, or between the two groups, for binary variables, as listed below arrow on each plot), BMI; body mass index (z-score). Breastfeeding is presented as the HR per three-month increase in breastfeeding duration within the first 10 months of life.

**Figure 2:**
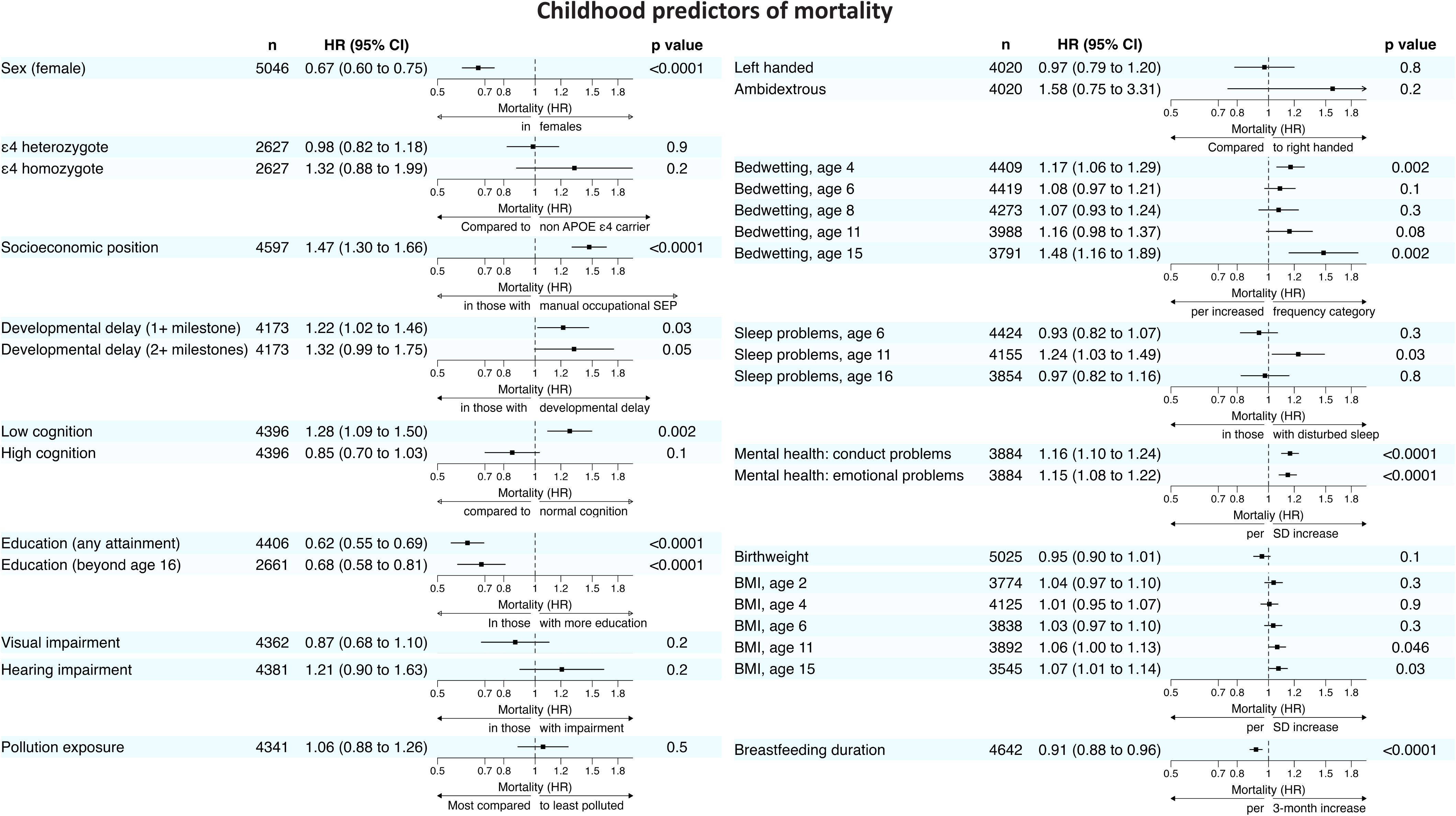
Childhood predictors of mortality. Forest plots demonstrating results of Cox proportional cause-specific hazards regression to examine the relationship between childhood factors and mortality by age 78, censoring for emigration, and adjusting for sex. Education beyond age 16 is compared only to those with any educational attainment up to age 16. HR; hazard ratio (per unit change, for continuous variables, or between the two groups, for binary variables, as listed below arrow on each plot), BMI; body mass index (z-score). Breastfeeding is presented as the HR per three-month increase in breastfeeding duration within the first 10 months of life.

**Table 1:** Participant characteristics.

| Characteristic | Overall<br>N = 5,046 | Dementia diagnosis? |  | Mortality? |  |
| --- | --- | --- | --- | --- | --- |
|  |  | No | Yes | No | Yes |
| Sex |  |  |  |  |  |
| Male | 2,636 (52%) | 2,574 (98%) | 62 (2.4%) | 1,887 (72%) | 749 (28%) |
| Female | 2,410 (48%) | 2,351 (98%) | 59 (2.4%) | 1,916 (80%) | 494 (20%) |
| Social class, age 11<sup>1</sup> |  |  |  |  |  |
| Non-manual | 1,895 (41%) | 1,847 (97%) | 48 (2.5%) | 1,507 (80%) | 388 (20%) |
| Manual | 2,702 (59%) | 2,633 (97%) | 69 (2.6%) | 1,928 (71%) | 774 (29%) |
| APOE ε4 allele count |  |  |  |  |  |
| Non-carrier | 1,841 (70%) | 1,795 (98%) | 46 (2.5%) | 1,427 (78%) | 414 (22%) |
| Heterozygous | 702 (27%) | 675 (96%) | 27 (3.8%) | 544 (77%) | 158 (23%) |
| Homozygous | 84 (3.2%) | 76 (90%) | 8 (9.5%) | 60 (71%) | 24 (29%) |
| Developmental delay |  |  |  |  |  |
| None | 3,725 (89%) | 3,643 (98%) | 82 (2.2%) | 2,808 (75%) | 917 (25%) |
| ≥1 milestone | 448 (11%) | 431 (96%) | 17 (3.8%) | 314 (70%) | 134 (30%) |
| ≥2 milestones | 162 (3.9%) | 153 (94%) | 9 (5.6%) | 111 (69%) | 51 (31%) |
| Educational attainment |  |  |  |  |  |
| None | 1,745 (40%) | 1,693 (97%) | 52 (3.0%) | 1,178 (68%) | 567 (32%) |
| Any | 2,661 (60%) | 2,602 (98%) | 59 (2.2%) | 2,093 (79%) | 568 (21%) |
| Beyond age 16 | 1,447 (33%) | 1,415 (98%) | 32 (2.2%) | 1,172 (81%) | 275 (19%) |
| Cognitive ability, age 8<sup>2</sup> |  |  |  |  |  |
| Normal | 3,244 (74%) | 3,175 (98%) | 69 (2.1%) | 2,433 (75%) | 811 (25%) |
| Low | 600 (14%) | 571 (95%) | 29 (4.8%) | 407 (68%) | 193 (32%) |
| High | 552 (13%) | 540 (98%) | 12 (2.2%) | 434 (79%) | 118 (21%) |
| Birthweight, kg | 3.40 (0.51) | 3.40 (0.51) | 3.40 (0.61) | 3.40 (0.51) | 3.39 (0.53) |
| BMI, kg/m<sup>2</sup> |  |  |  |  |  |
| Age 2 | 17.85 (2.51) | 17.85 (2.51) | 17.91 (2.40) | 17.82 (2.49) | 17.95 (2.55) |
| Age 4 | 16.18 (1.67) | 16.18 (1.67) | 16.19 (1.40) | 16.17 (1.67) | 16.22 (1.64) |
| Age 6 | 15.81 (1.38) | 15.81 (1.39) | 15.79 (1.34) | 15.79 (1.37) | 15.86 (1.43) |
| Age 11 | 17.37 (2.40) | 17.36 (2.40) | 17.92 (2.41) | 17.33 (2.35) | 17.50 (2.52) |
| Age 15 | 20.10 (2.78) | 20.09 (2.75) | 20.69 (3.51) | 20.06 (2.69) | 20.21 (3.02) |
| Handedness, age 11 |  |  |  |  |  |
| Right handed | 3,630 (90%) | 3,551 (98%) | 79 (2.2%) | 2,701 (74%) | 929 (26%) |
| Left handed | 372 (9.3%) | 354 (95%) | 18 (4.8%) | 277 (74%) | 95 (26%) |
| Ambidextrous | 18 (0.4%) | 16 (89%) | 2 (11%) | 11 (61%) | 7 (39%) |
| Nocturnal enuresis |  |  |  |  |  |
| Age 4 none | 3,902 (89%) | 3,813 (98%) | 89 (2.3%) | 2,944 (75%) | 958 (25%) |
| any | 507 (11%) | 486 (96%) | 21 (4.1%) | 350 (69%) | 157 (31%) |
| Age 6 none | 4,010 (91%) | 3,924 (98%) | 86 (2.1%) | 3,009 (75%) | 1,001 (25%) |
| any | 409 (9.3%) | 388 (95%) | 21 (5.1%) | 287 (70%) | 122 (30%) |
| Age 8 none | 4,025 (94%) | 3,933 (98%) | 92 (2.3%) | 3,008 (75%) | 1,017 (25%) |
| any | 248 (5.8%) | 234 (94%) | 14 (5.6%) | 174 (70%) | 74 (30%) |
| Age 11 none | 3,789 (95%) | 3,702 (98%) | 87 (2.3%) | 2,822 (74%) | 967 (26%) |
| any | 199 (5.0%) | 189 (95%) | 10 (5.0%) | 136 (68%) | 63 (32%) |
| Age 15 none | 3,716 (98%) | 3,628 (98%) | 88 (2.4%) | 2,776 (75%) | 940 (25%) |
| any | 75 (2.0%) | 71 (95%) | 4 (5.3%) | 44 (59%) | 31 (41%) |
| Breastfeeding, months | 4.6 (4.0) | 4.6 (4.0) | 4.5 (3.9) | 4.7 (4.0) | 4.1 (4.0) |
| Pollution exposure, up to age | 0.50 (0.33) | 0.50 (0.33) | 0.57 (0.33) | 0.50 (0.33) | 0.50 (0.33) |
| Mental health, age 13-15 |  |  |  |  |  |
| Conduct problems, z-score | 0.00 (1.00) | 0.00 (0.99) | 0.12 (1.19) | -0.05 (0.98) | 0.14 (1.03) |
| Emotional problems, z- | 0.00 (1.00) | 0.00 (1.00) | 0.37 (1.13) | -0.03 (0.97) | 0.11 (1.06) |
| Sleep disturbance |  |  |  |  |  |
| Age 6 none | 3,194 (72%) | 3,121 (98%) | 73 (2.3%) | 2,373 (74%) | 821 (26%) |
| any | 1,230 (28%) | 1,195 (97%) | 35 (2.8%) | 929 (76%) | 301 (24%) |
| Age 11 none | 3,738 (90%) | 3,655 (98%) | 83 (2.2%) | 2,796 (75%) | 942 (25%) |
| any | 417 (10%) | 402 (96%) | 15 (3.6%) | 291 (70%) | 126 (30%) |
| Age 16 none | 3,190 (83%) | 3,118 (98%) | 72 (2.3%) | 2,367 (74%) | 823 (26%) |
| any | 664 (17%) | 645 (97%) | 19 (2.9%) | 504 (76%) | 160 (24%) |
| Vision, age 6-15 |  |  |  |  |  |
| Normal | 4,051 (93%) | 3,952 (98%) | 99 (2.4%) | 3,011 (74%) | 1,040 (26%) |
| Impaired | 311 (7.1%) | 303 (97%) | 8 (2.6%) | 240 (77%) | 71 (23%) |
| Hearing, age 7-15 |  |  |  |  |  |
| Normal | 4,242 (97%) | 4,136 (98%) | 106 (2.5%) | 3,164 (75%) | 1,078 (25%) |
| Impaired | 139 (3.2%) | 134 (96%) | 5 (3.6%) | 94 (68%) | 45 (32%) |

Participant characteristics within the NSHD cohort with dementia/mortality data available. Values represent mean (standard deviation), and count (% of row for each diagnosis, column % for

### Childhood predictors of dementia

Associations between childhood factors and dementia by age 78 are shown in **Figure 1**.

Accounting for sex, increased dementia risk was seen in *APOE* L4 homozygotes (HR 4·19, 95% CI 1·98-8·88; *P*=0·0002), with a trend for increased risk in heterozygotes (HR 1·54, 95% CI 0·96-2·47; *P*=0·08). There was higher risk in those with delay in ≥1 (HR 1·81, 95% CI 1·07-3·05; *P*=0·03) or ≥2 milestones (HR 2·74, 95% CI 1·38-5·43; *P*=0·004); low cognition (HR 2·33, 95% CI 1·51-3·60; *P*=0·0001); higher pollution exposure (HR 2·05, 95% CI 1·16-3·64; *P*=0·01); left-handers (HR 2·22, 95% CI 1·33-3·71; *P*=0·002) or those classified as ambidextrous (HR 5·96, 95% CI 1·46-24·27; *P*=0·01); more frequent nocturnal enuresis at any time from age 4-15; more emotional problems (HR 1·46, 95% CI 1·21-1·78; *P*=0·0001); and higher BMI at age 11 (HR 1·21, 95% CI 1·02-1·45, *P*=0·03). There was a trend for higher dementia risk in those with sleep problems at age 11 (HR 1·72, 95% CI 0·99-2·98; *P*=0·05), and higher BMI at age 15 (HR 1·20, 95% CI 0·99-1·45; *P*=0·07).

There was decreased dementia risk in those with any educational attainment (HR 0·68, 95% CI 0·47-0·98; *P*=0·04), but no additional benefit of continuing education after age 16.

We found no significant relationship with sex, socioeconomic position, high cognition, birthweight, breastfeeding, visual, or hearing impairment.

Sensitivity analyses confirmed that associations with nocturnal enuresis remain after excluding (n=78) participants with nocturnal enuresis persisting at age 15 and that childhood handedness assessment was appropriate (**appendix [p7-8]**).

### Childhood predictors of mortality

Associations between childhood factors and mortality by age 78 are shown in **Figure 2**.

There was decreased mortality among females (HR 0·67, 95% CI 0·60-0·75, *P*<0·0001). Accounting for sex, mortality was lower among those with any education (HR 0·62, 95% CI 0·55-0·69, *P*<0·0001), with a further reduction in those continuing education beyond age 16 (HR 0·68, 95% CI 0·58-0·81, *P*<0·0001). Compared to those with normal cognition, individuals with low cognition had increased mortality (HR 1·23, 95% CI 1·09-1·50, *P*=0·002), those with high cognition had a trend for decreased mortality (HR 0·85, 0·70-1·03, *P*=0·1). Longer breastfeeding duration was associated with decreased mortality (HR 0·91/3-months, 95% CI 0·88-0·96, *P*<0·0001).

Lower socioeconomic position (HR 1·47, 95% CI 1·30-1·66, *P*<0·0001), delay in attaining ≥1 developmental milestones (HR 1·22, 95% CI 1·02-1·46, *P*=0·03), nocturnal enuresis at age 4 (HR 1·17, 95% CI 1·06-1·29, *P*=0·002) and age 15 (HR 1·48, 95% CI 1·16-1·89, *P*=0·002), sleep problems at age 11 (HR 1·24, 95% CI 1·03-1·49, *P*=0·03), conduct problems (HR 1·16, 95% CI 1·10-1·24, *P*<0·0001), emotional problems (HR 1·15, 95% CI 1·08-1·22, *P*<0·0001), and higher BMI at age 11 (HR 1·06, 95% CI 1·00-1·13, *P*=0·046) and age 15 (HR 1·07, 95% 1·01-1·14, *P*=0·03) were all associated with increased mortality by age 78.

There was no significant relationship with *APOE* L4 status, handedness, birthweight, visual, or hearing impairment.

These results are not due to infant/childhood mortality as no result was materially changed by excluding (n=261) participants who died or emigrated before age 18.

### Interactions with sex

#### Dementia

Analysis by sex revealed that relationships between higher BMI, left-handedness, and *APOE* L4 status and dementia risk were only present in females (**appendix [p11-13]**). Sensitivity analysis demonstrated that the relationship between BMI and dementia risk in females was materially unchanged after additionally controlling for age at menarche.

#### Mortality

The relationship between emotional problems and mortality risk was significantly stronger in females than in males (**appendix [p11-13]**). No other interactions with sex were observed.

### Independent predictors of mortality and dementia

We explored the independent contributions of all predictors with significant or near significant (*P*<0·1) relationship to dementia and mortality, using a multiply imputed dataset (n=5,046) to mitigate bias due to missing data.

#### Dementia

After stepwise elimination of predictors *P*>0·05, *APOE* L4 carriage, low cognition, pollution exposure, being left-handed/ambidextrous, disturbed sleep (age 11), more frequent nocturnal enuresis, higher BMI (age 11), and emotional problems were all significant independent predictors of dementia risk (**Figure 3A**). The interaction with sex in the relationship between both BMI and *APOE* ε4 status and dementia remained (**Figure 4A**), but the interaction between sex and handedness was no longer significant (*P*=0·1).

**Figure 3:**
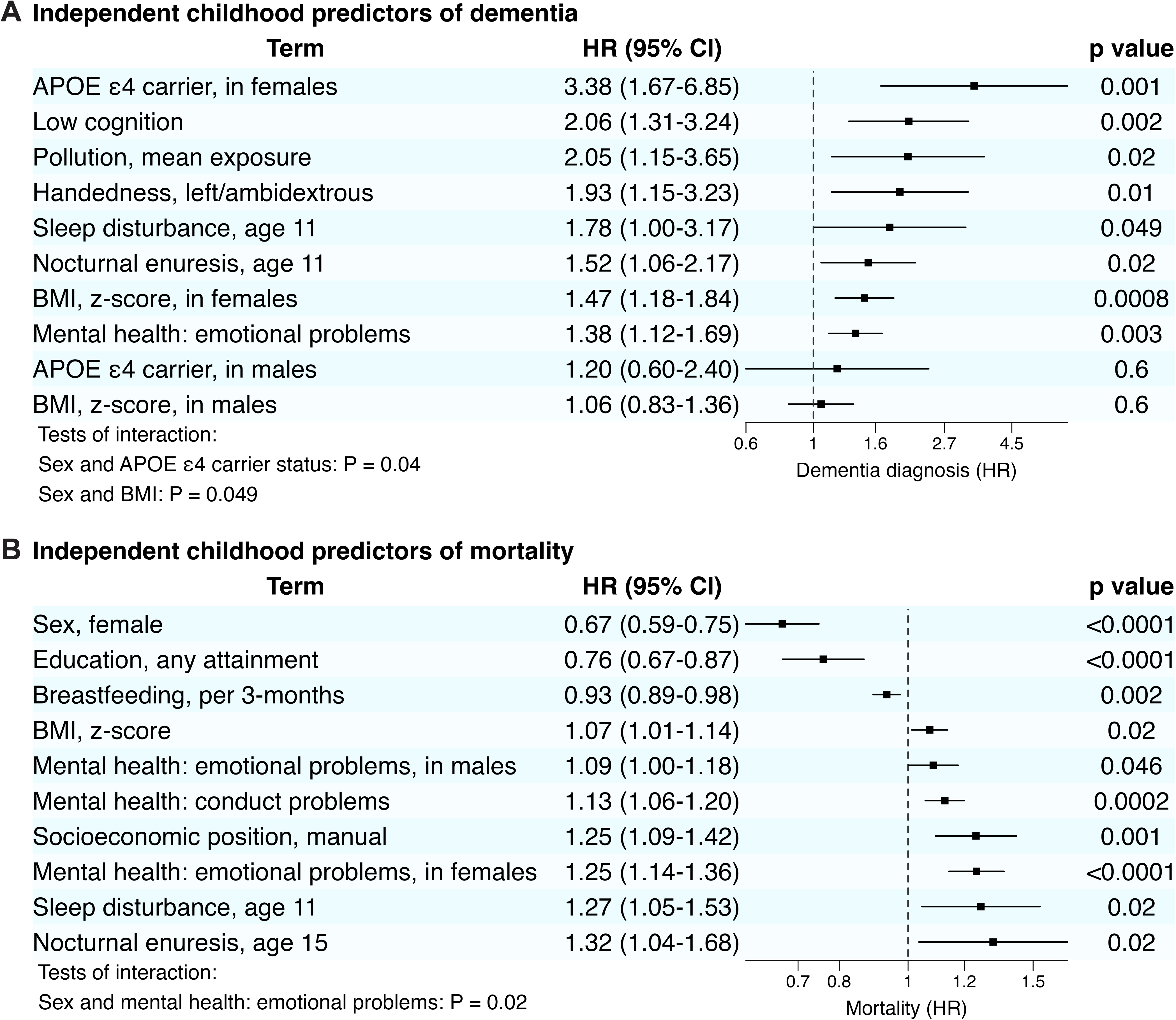
Independent childhood predictors of dementia and mortality. Forest plots demonstrating results of Cox proportional cause-specific hazards regression for dementia diagnosis by age 78 (**A**), censoring for emigration and for the competing risk of death, and for mortality by age 78 (**B**), censoring for emigration. Those factors with a significant or near significant (*P* < 0.1) relationship with mortality or dementia diagnosis in initial analyses (Figures 1-2) were selected for inclusion in initial models. Where a predictor was related to dementia or mortality at more than one time point, the closest to age 11 was selected. An overall effect of sex is not shown due to the presence of sex interactions. Backwards elimination was then used to iteratively exclude predictors that no longer remained significant (*P* < 0.05). To mitigate potential bias from missing data, all multivariable analyses were performed on multiply imputed datasets (n=5,046) using substantive model-compatible fully conditional specification (SMC-FCS). BMI; body mass index (z-score).

**Figure 4:**
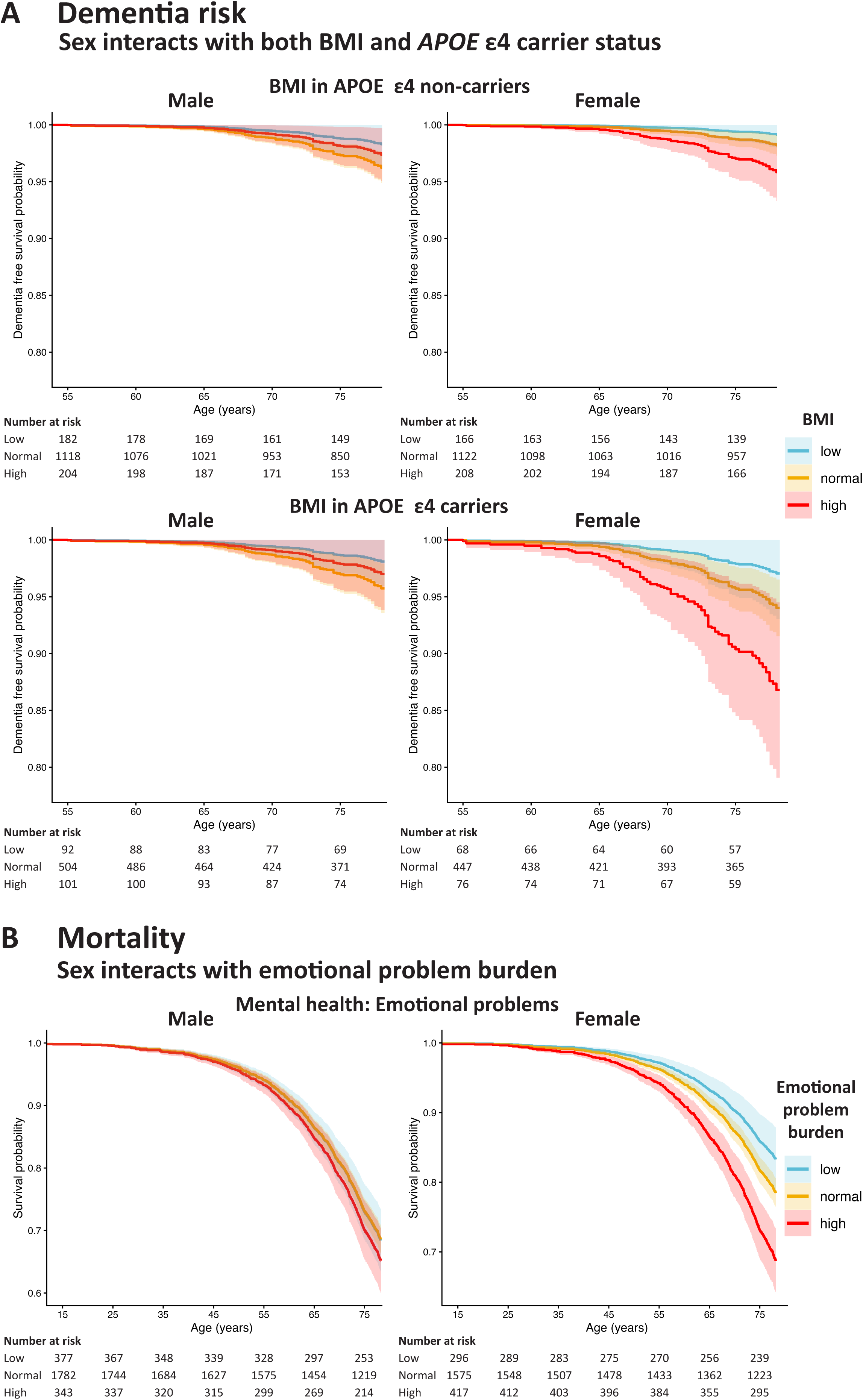
Independent childhood predictors of dementia and mortality; interactions with sex. **A**: Interaction between sex and both body mass index (BMI, z-score) and apolipoprotein E (*APOE*) L4 carrier status in predicting dementia risk. Adjusted cumulative survival plots show dementia-free survival, separately for males and females, stratified by *APOE* L4 carrier status, according to BMI, grouped here as low [>1 standard deviation (SD) below mean], normal [within 1 SD of mean], and high [>1 SD above mean]. Plots are adjusted for low cognitive ability, handedness, emotional problems (z-score), socioeconomic position, air pollution exposure, and *APOE* L4 carrier status, and includes interactions between sex and both BMI and *APOE* L4 carrier status (see Figure 3A). Plots for dementia diagnosis are truncated to start at the year of the first dementia diagnosis. **B**: Interaction between sex and emotional problems (z-score) in predicting mortality by age 78. Adjusted cumulative survival plots show mortality separately for males and females according to the burden of emotional problems, grouped here as low [>1 SD below mean], normal [within 1 SD of mean], and high [>1 SD above mean]. Plots are adjusted for sex, socioeconomic position, educational attainment, emotional problems, conduct problems, BMI (z-score, age 11), breastfeeding duration, nocturnal enuresis (age 15), sleep disturbance (age 11), and includes the interaction between sex and emotional problems (see Figure 3B).

#### Mortality

After stepwise elimination of predictors *P*>0·05, sex, education, breastfeeding, BMI (age 11), emotional problems, conduct problems, socioeconomic position, disturbed sleep (age 11), and nocturnal enuresis were all significant independent predictors of mortality (**Figure 3B**). Emotional problems were associated with increased mortality in both males and females, with a stronger relationship in females (**Figure 4B**).

Sensitivity analysis confirmed a monotonic relationship between dementia risk and BMI in females, and emotional problems, and between mortality and BMI, emotional problems (in females), and conduct problems (**appendix [p9-10]**).

### Population attributable fraction

To contextualise the potential contribution of these risk factors for dementia and mortality by age 78, we calculated PAF for all identified factors combined, and additionally excluding sex and *APOE* as clearly non-modifiable risks.

The total PAF due to childhood factors was 69·5% for dementia, and 56·7% for mortality. Excluding sex and *APOE*, the total PAF was 61·9% for dementia, and 43·5% for mortality. PAF for individual risks are listed in **appendix** [**p14**].

## Discussion

We demonstrate that a significant proportion of risk for both mortality and dementia by age 78 is predicted during childhood, by a wide range of factors including indices of compromised structural and functional brain development, genetic factors, and early-life exposures. These relationships, spanning eight decades, offer unique insights into factors that predispose to or protect against dementia and premature ageing, and highlight important opportunities for interventions.

### Childhood predictors of dementia

Independent childhood predictors of dementia include low cognition, pollution exposure, being left-handed/ambidextrous, sleep disturbance, nocturnal enuresis, and emotional problems; *APOE* L4 carriage and BMI were associated with dementia risk only in females.

The Lancet Commission include obesity and depression in midlife, and pollution exposure in late-life as risk factors for dementia. We find evidence that each of these associations begin in childhood. We also found some important differences; associations with childhood BMI were only present in females, while low BMI, a disputed risk factor for dementia during midlife,^2^ appears protective during childhood. Further study is required to understand these differences, noting that sex differences associated with BMI may relate to a range of factors including physical activity, metabolism, fat and muscle composition, and diet.^14^

We found education to be associated with lower dementia risk, but this association was no longer present after controlling for other childhood factors including cognition. Education is frequently reported to lower dementia risk but few studies can adjust for prospectively evaluated childhood cognition.^15^ Evidence from longitudinal adult cohorts finds that while education is associated with better cognitive ability, it does not protect against cognitive decline,^12^ suggesting that early-life factors which enable individuals to pursue education may drive associations between education and dementia risk. Our results support this conclusion, noting that educational opportunities in mid-twentieth-century Britain were closely linked to both socioeconomic position and cognitive ability.^15^

*APOE* ε4 allele carriage is the strongest genetic risk factor for Alzheimer’s disease (AD), the commonest cause of dementia, and it is unsurprising that we found it to predict dementia risk. Our finding that this association was only present in females aligns with growing evidence that female sex markedly enhances risk from the ε4 allele,^16^ although the relatively young age and all-cause dementia outcome, including non-AD dementias unrelated to *APOE* ε4 carriage, may also be a factor.

Nocturnal enuresis and being left-handed/ambidextrous were both associated with higher dementia risk, independent of other potentially related factors including disturbed sleep and symptoms of poor mental health. Neither of these factors are generally considered predictors of dementia, and while mechanisms for these associations are unclear, genetic factors and alterations in early brain development may both be important.^17^ Nocturnal enuresis is a highly genetic disorder, with 70% heritability in twin studies,^18^ while associations between left-handedness and genes encoding proteins implicated in both brain development and AD (*MAP2*, *MAPT*, *WNT3* and *MIC*) may underlie dementia risk in left-handers.^13^

Alterations in early brain development provide an alternative explanation for many associations between childhood factors and dementia risk, noting recent evidence that neurodevelopmental factors are associated earlier onset of AD.^17^ Increased dementia risk in those with low childhood cognitive ability is most readily explained by reduced cognitive reserve,^8^ but similar findings in those who are left-handed or ambidextrous suggest that otherwise benign variations in brain development may confer vulnerability to neurodegeneration.^13,17^ Nocturnal enuresis, generally believed to be a benign disorder, is associated with alterations in brain development.^9^ We find increased dementia risk in those with nocturnal enuresis from as early as 4 years old, independent of other factors including emotional problems, with higher risk in those who remain symptomatic later in childhood, suggesting that nocturnal enuresis may not always be benign, and in some cases may be a marker for vulnerability to late-life cognitive decline.

Sleep disturbance in late-life is associated with many forms of dementia including AD.^7^ There is evidence that this may play a causative role in the neurodegenerative process, potentially due to reduced sleep dependent clearance of pathological proteins from the CNS, but the association may also reflect reverse causation, occurring as a manifestation of very early disease.^2,7^ Associations between sleep disturbance at age 11 and dementia risk cannot be explained by reverse causation, and while disturbed sleep may reflect emotional problems, the association remained after controlling for a range of mental health metrics. While mechanisms explaining this association remain unclear, similar to nocturnal enuresis, sleep disturbances may be a manifestation of aberrant circadian control which signify or confer vulnerably to cognitive disease later in life.

### Childhood predictors of mortality

Independent childhood predictors of mortality include male sex, low education, less breastfeeding, higher BMI, less privileged socioeconomic position, sleep disturbance, nocturnal enuresis, and a greater burden of emotional and conduct problems.

Lower mortality risk in females,^19^ those with more education,^20^ and those from privileged socioeconomic backgrounds are well established.^21^ We additionally found that associations between high BMI, poor mental health, and disturbed sleep and increased mortality, known to be present in adulthood,^3,5,6^ all begin in childhood.

Unlike adults, in whom low BMI, often a feature of severe chronic disease, is associated with increased mortality,^6^ those with low childhood BMI had lower mortality risk by age 78. Early postwar Britain was characterized by a relatively high standard of nutrition and it is unlikely that this finding is related to malnutrition, which is particularly harmful during childhood.^22^ It may relate to differences in metabolism, proteomic aging, or lower rates of adult obesity in those with lower childhood BMI,^23,24^ while associations between low-calorie diets and increased longevity observed in adult populations may also be relevant.^25^

We found breastfeeding to be associated with lower mortality after childhood, up to age 78, independent of a range determinants of health including parental social class, in line with findings from large retrospective analyses.^10^ Advantages of breastfeeding are generally attributed to its interactional nature and the unique properties of breastmilk, conferring benefits including improved nutrition, lower rates of infection, improved brain development and cognition.^10^

We found that higher childhood cognitive ability associates with lower mortality, but this relationship was no longer present when other childhood factors, including socioeconomic position and education are taken into account, suggesting that previously described associations between cognition and mortality may be driven by other factors.^26^

Associations between nocturnal enuresis and increased mortality risk were unexpected. Further research should explore whether this association is underpinned by genetic factors, which may co-associate with other pathways which increase risk.^18^ Interactions between nocturnal enuresis, sleep, and mental health may be important, however, nocturnal enuresis independently predicted mortality in models including these factors.

### Population attributable fraction

Our exploratory post-hoc analyses suggests that taken together, factors present in the childhood period are associated with more than half the risk of both dementia and mortality by age 78.

When including all identified risk factors in the PAF calculation we found that 69.5% of dementia risk and 56.7% of mortality could be explained; reducing slightly to 61.9% and 43.5% when excluding *APOE* status and sex respectively. Prior studies of potentially modifiable risks assessed predominantly during adult life report PAFs for dementia ranging from 24-67%.^27^ The relatively high PAF we report may in part relate to the significantly longer exposure spanning nearly eight decades, the ability of birth cohorts to capture underrepresented populations who may have a greater risk burden, and the backwards elimination approach which may result in overrepresentation of features with large effect sizes. Even where risk factors are modifiable, it is unclear to what extent this may reduce or delay risk, while modification of risks to low levels, as assumed by PAF, may not be possible. Thus, while caution is required when interpreting the magnitude of these results, they nevertheless suggest that influences present in childhood contribute a non-trivial proportion of risk to late-life brain and general health.

### Strengths and limitations

This study has the longest continuous prospective follow-up of any cohort in the world. Participants are the same age with extensive prospective data collection enabling rigorous correction for life course determinants of general and brain health. National collection of diagnostic and mortality data provides a robust reporting mechanism but is limited to dementia diagnoses captured by HES within the NHS in England, and likely under-reports diagnoses, particularly at early stages.^28^

While we were able to robustly adjust for a range of childhood determinants of health and disease, it is unknown to what extent reported associations were due to the childhood exposures themselves or their effects or associations with other factors into adulthood.

Age 78 remains relatively young for dementia diagnosis. The number of dementia diagnoses may prevent detection of more subtle associations, particularly where stratified by sex, while differing precision in ascertainment of childhood exposures may result in residual confounding. We present unadjusted *P*-values to preserve statistical power, acknowledging that significant findings should be validated in independent cohorts. Relatively poor correlation between clinical and neuropathological dementia diagnosis, and high rates of mixed pathology, necessitated the grouping together of all-cause dementia diagnoses.^29^ Finally, reflecting those born in Britain in 1946 the cohort is exclusively white and it is unclear if these findings are representative of other ethnic groups.

### Implications

Childhood factors associated with mortality and dementia each have a range of longer-term effects and associations with life course health and behaviours, initiating chains of risk extending into late-life.^8^ Some risk factors we identified are already established as risks in later-life and we can extend evidence back to childhood. For others, further study is required to examine causal pathways and identify whether modification can improve health outcomes.

Both dementia and mortality were predicted by higher BMI, sleep disturbance, nocturnal enuresis, and greater burden of emotional problems. There were also several important differences. Specific risk factors for dementia such as cognition and handedness suggest that subtle neurodevelopmental differences may be of particular importance in determining risk/resilience to dementia. Apart from sex, every childhood risk factor for mortality was potentially modifiable.

We found BMI to be associated with dementia and mortality only after the end of post-war food rationing (1954, age 8), prior to which the British population was subject to a limited but generally nutritious diet, irrespective of income, resulting in improved health outcomes,^22^ suggesting that childhood dietary interventions may improve long term brain and general health. Effective treatments for sleep disorders, mental health disorders, and nocturnal enuresis imply that these underdiagnosed risk factors for both dementia and mortality are potentially modifiable.^18,30^ Future research should endeavour to understand if these factors, when present in childhood, are symptoms or cause of vulnerability to poorer late-life brain and general health, and whether modification can improve outcomes. Should these associations be due to alterations in early brain development, it would further support the importance of antenatal and perinatal care for brain and general health.

### Conclusion

We show that features from the first years of life predict a significant proportion of mortality and dementia risk into the eighth decade. While some of these risks are genetically determined or may relate to subtle differences in brain development, many are potentially modifiable. Potentially modifiable risk factors for brain and general health including education, mental health, sleep, nocturnal enuresis, socioeconomic position, breastfeeding, BMI, and pollution exposure offer crucial opportunities to improve health outcomes over the human lifetime and highlight the potential for targeted interventions during childhood.

## Acknowledgements

We are very grateful to those study members who helped in the design of the study through focus groups, and to the participants both for their contributions to Insight 46 and for their commitments to research over the past eight decades. We are grateful to the radiographers and nuclear medicine physicians at the University College London Institute of Nuclear Medicine and to the staff at the Leonard Wolfson Experimental Neurology Centre at University College London. We also thank Dr Katie Malbon and Dr Delia Toomey for their valuable contributions to the interpretation of these findings.

## Funding

Insight 46 is funded by grants from Alzheimer’s Research UK (ARUK-PG2014-1946, ARUK-PG2017-1946), Alzheimer’s Association (SG-666374-UK BIRTH COHORT), The Wolfson Foundation (PR/ylr/18575), The Medical Research Council (MC-UU-12019/1, MC-UU-12019/3), Weston Brain Institute, and Brain Research UK (UCC14191).

J.K-R. is supported by a Medical Research Council Clinical Research Training Fellowship (MR/Y009452/1).

J.M.S. is a National Institute for Health Research (NIHR) Senior Investigator and acknowledges the support of the NIHR University College London Hospitals Biomedical Research Centre and the UCL Centre of Research Excellence, an initiative funded by British Heart Foundation (RE/24/130013). This work is supported by the UK Dementia Research Institute through UK DRI Ltd, principally funded by the Medical Research Council. He has grant funding from Alzheimer’s Research UK, Brain Research UK, Weston Brain Institute, Medical Research Council, British Heart Foundation, Wolfson Foundation, and Alzheimer’s Association. This work uses data provided by study participants or patients and collected through clinical studies or as part of their care and support.

J.W.G. is supported by an Alzheimer’s Research UK (ARUK) Clinical Research Training Fellowship (ARUK-CRTF2023B-001).

Data collection for the cohort and salary support for N.C. was provided by the Medical Research Council (MC_UU_12019/1).

## Competing interests

J.M.S has consulted for Roche, Eli Lilly, Biogen, AVID, Merck and GE, and received royalties from Oxford University Press and Henry Stewart Talks. He serves on scientific advisory boards for Receptive Bio and Alamar Biosciences. He is Chief Medical Officer for Alzheimer’s Research UK. NC serves on data monitoring and safety committees of trials sponsored by AstraZeneca.

## Author contributions

J.K-R and J.M.S. conceptualized the study and are the guarantors. J.K-R. led study design, literature search, data analysis and visualisation, and wrote the original draft.

J.M.S. supervised the study. J.N provided statistical input. S-N.J, J.G., N.C., and M.R. provided analytical input. All authors reviewed and critically revised the manuscript for intellectual content. The corresponding author attests that all listed authors meet authorship criteria and that no others meeting the criteria have been omitted. All authors read and approved the final version of the manuscript. J.K-R and J.N. have accessed and verified the underlying data reported in the manuscript.

## Role of the Funder/Sponsor

The funders and sponsors had no role in the design and conduct of the study; collection, management, analysis, and interpretation of the data; preparation, review, or approval of the manuscript; and decision to submit the manuscript for publication

## Data availability

The NSHD data-sharing policy is available on the NSHD Data Sharing website (https://nshd.mrc.ac.uk/data-sharing). Anonymised data and a data dictionary are available to researchers upon request to the NSHD Data Sharing Committee via a standard application procedure. Childhood variables used for analysis are collated in basket joshkrZZjiolqt on the Condor data sharing platform. Mortality and Hospital Episode Statistics data can be requested from the UK Longitudinal Linkage Collaboration (https://ukllc.ac.uk/).

## Transparency

The corresponding authors affirm that the manuscript is an honest, accurate, and transparent account of the study being reported; that no important aspects of the study have been omitted; and that any discrepancies from the study as planned have been explained.

## Notes

### Author Declarations

National Research Ethics Service Committee London (REC reference:14/LO/1173) gave ethical approval for this work.

## References

1. Office for National Statistics. Leading causes of death, UK: 2001 to 2018. 2020.

2. Livingston G, Huntley J, Liu KY, et al. Dementia prevention, intervention, and care: 2024 report of the Lancet standing Commission. Lancet 2024; 404(10452): 572–628.

3. Machado MO, Veronese N, Sanches M, et al. The association of depression and all-cause and cause-specific mortality: an umbrella review of systematic reviews and meta-analyses. BMC Med 2018; 16(1): 112.

4. Tan BKJ, Ng FYC, Song H, Tan NKW, Ng LS, Loh WS. Associations of Hearing Loss and Dual Sensory Loss With Mortality: A Systematic Review, Meta-analysis, and Meta-regression of 26 Observational Studies With 1L213L756 Participants. JAMA Otolaryngol Head Neck Surg 2022; 148(3): 220–34.

5. Cappuccio FP, D’Elia L, Strazzullo P, Miller MA. Sleep duration and all-cause mortality: a systematic review and meta-analysis of prospective studies. Sleep 2010; 33(5): 585–92.

6. Nowak MM, Niemczyk M, Golebiewski S, Paczek L. Impact of Body Mass Index on All-Cause Mortality in Adults: A Systematic Review and Meta-Analysis. J Clin Med 2024; 13(8).

7. Wang C, Holtzman DM. Bidirectional relationship between sleep and Alzheimer’s disease: role of amyloid, tau, and other factors. Neuropsychopharmacology 2020; 45(1): 104–20.

8. James S-N, Schott JM, Ben-Shlomo Y. A life course approach to neurodegeneration. In: Kuh D, Susser E, Blodgett JM, Ben-Shlomo Y, eds. A Life Course Approach to the Epidemiology of Chronic Diseases and Ageing: Oxford University Press; 2025: 209–28.

9. Lei D, Ma J, Shen X, et al. Changes in the brain microstructure of children with primary monosymptomatic nocturnal enuresis: a diffusion tensor imaging study. PLoS One 2012; 7(2): e31023.

10. Wang X, Yan M, Zhang Y, et al. Breastfeeding in infancy and mortality in middle and late adulthood: A prospective cohort study and meta-analysis. J Intern Med 2023; 293(5): 624–35.

11. Wadsworth M, Kuh D, Richards M, Hardy R. Cohort Profile: The 1946 National Birth Cohort (MRC National Survey of Health and Development). Int J Epidemiol 2006; 35(1): 49–54.

12. Fjell AM, Rogeberg O, Sorensen O, et al. Reevaluating the role of education on cognitive decline and brain aging in longitudinal cohorts across 33 Western countries. Nat Med 2025; 31(9): 2967–76.

13. Wiberg A, Ng M, Al Omran Y, et al. Handedness, language areas and neuropsychiatric diseases: insights from brain imaging and genetics. Brain 2019; 142(10): 2938–47.

14. Campbell MK. Biological, environmental, and social influences on childhood obesity. Pediatr Res 2016; 79(1-2): 205–11.

15. Richards M, Sacker A. Lifetime antecedents of cognitive reserve. J Clin Exp Neuropsychol 2003; 25(5): 614–24.

16. Ungar L, Altmann A, Greicius MD. Apolipoprotein E, gender, and Alzheimer’s disease: an overlooked, but potent and promising interaction. Brain Imaging Behav 2014; 8(2): 262–73.

17. Miller ZA, Ossenkoppele R, Graff-Radford NR, et al. Neurodevelopment and neural environment inform Alzheimer’s disease age at onset and phenotype. Alzheimers Dement 2025; 21(9): e70668.

18. Radmayr CD, HS; Gali, EB; Haid, B; Hoen, L; Kocvara, R; Nijman, RJM; Silay, MS; Stein, R; Tekgül, S. EAU Guidelines on Paediatric Urology: Monosymptomatic nocturnal enuresis. EAU Guidelines Office, Arnhem, The Netherlands., 2025.

19. Wu YT, Niubo AS, Daskalopoulou C, et al. Sex differences in mortality: results from a population-based study of 12 longitudinal cohorts. CMAJ 2021; 193(11): E361–E70.

20. Collaborators I-C. Effects of education on adult mortality: a global systematic review and meta-analysis. Lancet Public Health 2024; 9(3): e155–e65.

21. Kuh D, Hardy R, Langenberg C, Richards M, Wadsworth ME. Mortality in adults aged 26-54 years related to socioeconomic conditions in childhood and adulthood: post war birth cohort study. BMJ 2002; 325(7372): 1076–80.

22. Boudreau FG. Nutrition in war and peace. 1947. Milbank Q 2005; 83(4): 609-

23. Wang G, Wei D, Kebede Merid S, et al. BMI trajectories from birth to young adulthood associate with distinct cardiometabolic profiles. BMC Med 2024; 22(1): 510.

24. Groves JW, Bot VA, Ding DY, et al. [preprint] Eight decades of follow-up link life course exposures to proteomic organ ageing and longevity. medRxiv 2025: 2025.09.07.25335188.

25. Di Francesco A, Deighan AG, Litichevskiy L, et al. Dietary restriction impacts health and lifespan of genetically diverse mice. Nature 2024; 634(8034): 684–92.

26. Calvin CM, Batty GD, Der G, et al. Childhood intelligence in relation to major causes of death in 68 year follow-up: prospective population study. BMJ 2017; 357: j2708.

27. Stephan BCM, Cochrane L, Kafadar AH, et al. Population attributable fractions of modifiable risk factors for dementia: a systematic review and meta-analysis. Lancet Healthy Longev 2024; 5(6): e406–e21.

28. Brown A, Kirichek O, Balkwill A, et al. Comparison of dementia recorded in routinely collected hospital admission data in England with dementia recorded in primary care. Emerg Themes Epidemiol 2016; 13: 11.

29. Brunnstrom H, Englund E. Clinicopathological concordance in dementia diagnostics. Am J Geriatr Psychiatry 2009; 17(8): 664–70.

30. Hornsey SJ, Gosling CJ, Jurek L, et al. Umbrella Review and Meta-Analysis: The Efficacy of Nonpharmacological Interventions for Sleep Disturbances in Children and Adolescents. J Am Acad Child Adolesc Psychiatry 2025; 64(3): 329–45.

