## appendix for "Childhood predictors of dementia and mortality: 78 year follow-up of the 1946 British Birth Cohort"

**Supplementary appendix**

### Appendix 1: Childhood predictors. Data collection and variable definitions:

Sex was ascertained at birth. Gender data is not available.

*APOE* ɛ4 carrier status was determined from blood sampling at age 53, classified as ‘non carrier’, ‘heterozygous’ or ‘homozygous’ for the *APOE* ɛ4 allele.^1,2^

Educational attainment was defined as the highest qualification by age 26 based upon the Burnham Scale, categorised as none, any education (including vocational, sub-GCSE, GCSE level and above), and education beyond age 16 (A-level, degree level, and postgraduate education).^3^ In order to capture educational attainment of all durations including part time qualifications, attainment was ascertained at age 26, and may have included some participants who commenced education outside of the childhood years.

Paternal social class was determined when the participant was aged 11 (or 15 or 4 where unavailable) using the Office of Population Censuses and Surveys classification of the father’s occupation as non-manual (skilled non-manual, intermediate, and professional) and manual (unskilled, partly skilled, skilled manual).^4^

Cognition was assessed at age 8 combining four tests of verbal and non-verbal ability, and converted into a z-score standardised to the full available cohort.^3^ Where unavailable at 8, scores at age 11 or 15 were used. Individuals with cognitive scores less than or more than one standard deviation (SD) from the mean were defined as having low, and high cognition, respectively.

Breastfeeding duration during the first 10 months of life was ascertained during a structured interview with the participant’s mother by a health visitor when the participant was 2 years old.

Age (in months) individuals attained developmental milestones (sitting unaided, standing unaided, walking several steps without support and talking more than “mum”, “dad” or “nan”) was ascertained at interview with the participant’s mother by a health visitor at the age of 2 years. Developmental delay was defined as reaching each milestone later than 95% of the cohort and categorised as no delay, one or more delayed milestones, or two or more delayed milestones.

Body mass index (BMI) was calculated from height and weight measured at age 2, 4, 6, 11, and 15. z-scores were calculated separately for males and females from the cohort mean and SD at that age.^5^ Birthweight was converted to z-score using the same methodology.

Childhood mental health was assessed from teachers’ assessment of behaviour at age 13 and 15.^6,7^ We used previously derived measures based on factor analyses of scales to generate metrics of emotional problems (anxiety, timidity, fearfulness, diffidence and avoidance of attention) and conduct problems (unpunctuality, restlessness, truancy, daydreaming, indiscipline, disobedience and lying),^6,7^ which have previously been shown to be strongly predictive of later life mental health disorders.^6,8^ The mean score for the two ages was converted to a z-score based on the full cohort.

Nocturnal enuresis (bed wetting) was ascertained during maternal interview at ages 4, 6, 8, and with the school doctor at age 11, and 15, and classified as ‘never’, ‘occasionally’, ‘several nights per week’ and ‘always’. Ordinal responses were recoded (0–3) and treated as a continuous linear predictor to assess the presence of a dose-response relationship.

Individuals were classified as having sleep disturbance when the mother reported that a family member had to go to the child because of nightmares or disturbed sleep at least ‘occasionally’ in the prior year during an interview at ages 6 and 11, or where the study participant reported trouble with sleep at age 16.

Hearing was assessed by a doctor at age 7, 11 and 15. Those identified as having ‘poor’ or ‘very poor’ hearing at any age were classified as having hearing impairment.^7^

Visual acuity was assessed without glasses by a doctor using a Snellen chart at age 6, 7, 11, and 15. Vision was considered impaired where acuity of the better seeing eye was worse than 6/18 at any assessment.^9^

Air pollution at the child’s home address was estimated at birth and ages 2, 4, 6, 7, 8, 9, and 11, and categorised into four levels based upon local industry and coal consumption as previously described,^10^ from which the mean childhood exposure was estimated across all available years of data, and re-scaled from least (0) to most (1) exposed.

Handedness was determined by a doctor at age 11 as ‘right-handed’, ‘left-handed’, or ‘ambidextrous’. To confirm validity of childhood assessment of handedness, we compared this to detailed quantification of handedness using handedness questionnaire, which assesses hand preference for 12 different tasks using a numerical scale, performed in a subset of participants at age 70.^1,11^

Age at menarche was recorded during assessment by a doctor at age 15.^12^

### Appendix 2: Mortality and dementia data.

Hospital Episode Statistics (HES): This includes diagnoses at all NHS funded hospital admissions and outpatient appointments in England.^13^ In the absence of pathological confirmation, noting poor correlation between clinical and pathological diagnoses even in expert centres,^14^ we classified individuals as having dementia based on having one or more ICD codes F00, F01 F02, F03, G30, G31.^15^

### Appendix 3: Supplementary information on statistical approach

All analysis was performed in R (version 4·3·1).^16^

#### Cox regression

Cox proportional hazards regression was used to examine the relationship between childhood factors and risk of mortality and dementia diagnosis by age 78. Mortality was censored for age at emigration, while models examining dementia diagnosis were censored for the competing event of death and for age at emigration. The proportional hazards assumptions were assessed using Schoenfeld residual tests, and linearity of continuous predictors was assessed using plots of martingale residuals against the predictors.

For multivariate analyses, where a predictor was related to dementia or mortality risk at more than one time point, the measurement closest to age 11 was selected as this time point had the most consistent data availability.

#### Multiple imputation

To mitigate potential bias from missing data all multivariable analyses were performed on imputed datasets**.**

Multiple imputation of covariates used substantive model-compatible fully conditional speciﬁcation (SMC-FCS, 50 burn-in iterations, 50 imputations) in R to produce separate multiply imputed datasets for analyses of independent predictors of dementia risk and independent predictors of mortality.^17^ This approach imputes the missing data to be compatible with the Cox proportional cause-specific hazards substantive models for which the data will be used, and generally outperforms traditional multiple imputation approaches, particularly where interactions are present.^17^ We included the Nelson–Aalen marginal baseline cumulative hazard function for dementia diagnosis and mortality to further improve imputation results.^18^

Imputed datasets included data from all 5046 participants with mortality and dementia diagnosis data available. Dementia diagnosis, mortality and emigration outcomes were not imputed.

All factors showing a relationship with mortality at the *P* < 0·1 level were selected for inclusion in multivariate Cox proportional cause-specific hazards models, which was used as the basis for the substantive model and included all significant interactions with sex.

**Imputation of dataset for analysis of independent predictors of dementia diagnosis**

Substantive model type:	Competing risks

Substantive models (R code):

Surv(dementia_free_survival, event_competing_risks == 1) ~ sex + education_any + polmean + cog_binary + bmi57u_z + han57_binary + inefa + apoe_status + niw57 + nig5657_binary + any_developmental_delay + sex:bmi57u_z + sex:apoe_status + sex:han57_binary)

Surv(dementia_free_survival, event_competing_risks == 2) ~ sex + education_any + polmean + cog_binary + bmi57u_z + han57_binary + inefa + apoe_status + niw57 + nig5657_binary + any_developmental_delay + sex:bmi57u_z + sex:apoe_status + sex:han57_binary)

**Imputation of dataset for analysis of independent predictors of mortality**

Substantive model type:	Cox regression

Substantive model (R code):

Surv(mortality_survival, event_mortality) ~ sex + chsc_binary + any_developmental_delay + cog_trinary + education_any + niw61 + nig5657_binary + inefa + exefa + bmi61u_z + bre + inefa:sex")

#### Variables included in imputation models and imputation method:

Models included all variables from the substantive models in addition to auxiliary variables predictive of missing covariate data or missingness.

Dementia diagnoses depend upon reporting via hospital episode statistics but are treated as fully observed^1^ for the purpose of the imputation. Following imputation, continuous data were constrained to match the limits of the original measurement range,^2^ or converted to z-score based upon the mean and standard deviation within each imputed dataset,^3^ as appropriate. Missing represents the number pf participants with missing data for the listed variable, prior to multiple imputation. Where possible, variable names are as listed on the Condor data sharing platform, unless derived where descriptive titles are provided.

| **Variable name** | **Description** | **Imputation method** | **Imputed dataset** | | **Missing**  (of 5046) |
| --- | --- | --- | --- | --- | --- |
|  |  |  | **Dementia** | **Mortality** |  |
| event_competing_risks | Event marker, none (0), dementia (1), death (2). | Fully observed^1^ | ● |  | 0 |
| event_dementia | Event marker, dementia diagnosis. | Fully observed^1^ | ● |  | 0 |
| dementia_free_survival | Age (years) at dementia diagnosis/death/emigration. | Fully observed^1^ | ● |  | 0 |
| event_mortality | Event marker, death. | Fully observed |  | ● | 0 |
| mortality_survival | Age (years) at death/emigration. | Fully observed |  | ● | 0 |
| sex | Sex. | Fully observed | ● | ● | 0 |
| hazard_dementia | Cumulative hazard rate (Nelson-Aalen estimator) for dementia. | Fully observed | ● | ● | 0 |
| hazard_death | Cumulative hazard rate (Nelson-Aalen estimator) for death. | Fully observed | ● | ● | 0 |
| educational_attainment | Educational attainment, on Burnham scale (1-5). | Proportional odds regression | ● | ● | 640 |
| education_any | Any educational attainment. | Passively imputed: educational_attainment > 1 | ● | ● | 640 |
| chsc_binary | Child social class. | Bias reduced logistic regression | ● | ● | 449 |
| apoe4_count | APOE ε4 allele count. | Proportional odds regression | ● | ● | 2419 |
| apoe_status | APOE ε4 carrier. | Passively imputed: apoe_count > 0 | ● | ● | 2419 |
| any_developmental_delay | Nonor any developmental delay. | Bias reduced logistic regression | ● | ● | 873 |
| multiple_developmental_delays | Developmental delay in <2 or ≥2 milestones. | Bias reduced logistic regression | ● | ● | 873 |
| bmi48u_z | BMI, age 2 (z-score). | Linear regression | ● | ● | 1273 |
| bmi50u_z | BMI, age 4 (z-score). | Linear regression | ● | ● | 921 |
| bmi52u_z | BMI, age 6 (z-score). | Linear regression | ● | ● | 1208 |
| bmi53u_z | BMI, age 7 (z-score). | Linear regression | ● | ● | 1121 |
| bmi57u_z | BMI, age 11 (z-score). | Linear regression | ● | ● | 1154 |
| bmi61u_z | BMI, age 15 (z-score). | Linear regression | ● | ● | 1501 |
| cog_binary | Cognition, grouped into low (z-score <-1), normal/high (z-score ≥-1) | Passively imputed: COG81115 <-1 = low | ● |  | 650 |
| cog_trinary | Cognition, grouped into low (z-score <-1), normal (z-score -1 to 1) and high (z-score >1) | Passively imputed: COG81115 <-1 = low, -1 to 1 = normal, >1 = high |  | ● | 650 |
| COG81115 | Cognition at age 8, or age 11/15 where not available, z-score. | Linear regression | ● | ● | 650 |
| han57_binary | Right-handed or left-handed/ambidextrous | Bias reduced logistic regression | ● | ● | 1026 |
| bre^2^ | Breastfeeding duration (months) | Linear regression | ● | ● | 404 |
| niw50^2^ | Nocturnal enuresis frequency age 4. | Linear regression | ● | ● | 637 |
| niw52^2^ | Nocturnal enuresis frequency age 6. | Linear regression | ● | ● | 627 |
| niw54^2^ | Nocturnal enuresis frequency age 8. | Linear regression | ● | ● | 773 |
| niw55^2^ | Nocturnal enuresis frequency age 9. | Linear regression | ● | ● | 823 |
| niw57^2^ | Nocturnal enuresis frequency age 11. | Linear regression | ● | ● | 1058 |
| niw61^2^ | Nocturnal enuresis frequency age 15. | Linear regression | ● | ● | 1255 |
| nig5152 | Sleep disturbance frequency, age 6 | Linear regression | ● | ● | 622 |
| nig5657 | Sleep disturbance frequency, age 11 | Linear regression | ● | ● | 891 |
| slcna | Sleep disturbance, age 16 | Bias reduced logistic regression | ● | ● | 1192 |
| nig5152_binary | Sleep disturbance, age 6 | Passively imputed: nig5152 > 0.5 (equivalent to rounding) | ● | ● | 622 |
| nig5657_binary | Sleep disturbance, age 11 | Passively imputed: nig5657 > 0.5 (equivalent to rounding) | ● | ● | 891 |
| polmean | Pollution exposure (birth to age 11) | Linear regression | ● | ● | 705 |
| ex13efa | C\onduct problems age 13. | Linear regression | ● | ● | 984 |
| ex15efa | Conduct problems age 15. | Linear regression | ● | ● | 1040 |
| in13efa | Emotional problems age 13. | Linear regression | ● | ● | 984 |
| in15efa | Emotional problems age 15. | Linear regression | ● | ● | 1040 |
| exefa^3^ | Conduct problems age 13-15. | Passively imputed: ex13efa + ex15efa | ● | ● | 1162 |
| inefa^3^ | Emotional problems age 13-15. | Passively imputed: in13efa + in15efa | ● | ● | 1162 |

#### Population attributable fraction

The population attributable fraction (PAF) was calculated from the cox models under the counterfactual scenario where risk factors were not present.

$$PAF=\frac{P_{observed}- P_{expected}}{P_{observed}}$$

Where $P_{observed}$ represents the observed risk, and $P_{expected}$ represents the estimated risk under the counterfactual scenario.

We calculated both the total PAF; the estimated scenario where all risk factors were not present, as well as the individual PAF; where each single risk factor was not present, separately. In the case of continuous exposures, where zero exposure can be nonsensical or impractical (such as body mass index (BMI)), we estimated the effect of shifting the exposure value to the closest value that is within the lowest 1/3^rd^ of risk.^19^ All PAF calculations were pooled across multiply imputed datasets.

We included all risk factors, including those that are not modifiable, in this calculation. This enables a fuller understanding of entire spectrum of risk, noting that the extent to which many of these risk factors are modifiable remains very uncertain.

### Appendix 4: Data visualisation

Adjusted survival plots were predicted from cox regression models using a G-formula approach in the adjustedCurves package.^20^ To enable visualisation of the effect of different levels of continuous predictors, survival curves were predicted grouping z-scores into low (>1 SD below the mean), normal (within ±1 SD of the mean), and high (>1 SD above the mean). Survival probabilities are averaged across multiply imputed datasets according to Rubins Rules.

### Appendix 5: Sensitivity analyses- methods

Childhood mortality: A sensitivity analysis excluded all participants who died or emigrated prior to age 18 from mortality analyses, to exclude childhood mortality as the driver for significant results.

Nocturnal enuresis: To confirm that associations with nocturnal enuresis remain in those who stop bedwetting by the end of childhood, a sensitivity analysis examined associations with nocturnal enuresis excluding those with nocturnal enuresis persisting at age 15.

Age at menarche: To examine whether associations between BMI and health outcomes were driven by age of puberty, a sensitivity analysis additionally adjusted for age at menarche.

Linearity assumption: Sensitivity analyses were performed for significant results for continuous predictors represented as z-scores to examine whether the linearity assumption was appropriate. For this, z-scores were converted into a three-level factor variable: low (>1 SD below the mean), normal (within ±1 SD of the mean), and high (>1 SD above the mean). Cox regression then examined the relationship between these factors and the outcome.

### Appendix 6: Sensitivity analyses- results

#### Nocturnal enuresis

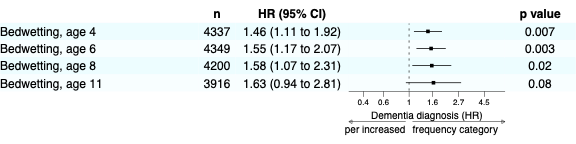
Forest plots demonstrating results of Cox proportional cause-specific hazards regression to examine the relationship between childhood nocturnal enuresis during childhood and dementia diagnosis by age 78, censoring for emigration the competing risk of death, and adjusting for sex, and excluding 78 participants who had nocturnal enuresis persisting at age 15.

#### Handedness

Childhood assessment of handedness determined by a doctor at age 11 was compared to detailed handedness assessment in a subset (n=472) of participants at age 70 using the handedness questionnaire. The handedness questionnaire examines hand preference across 12 tasks, giving a total score from -24 (strong left-handed) to +24 (strong right handed), with scores ≤-9 classified as left handed, and ≥9 as right handed.^11^ When assessments at age 11 and 70 are compared, 97·4% of those assessed as right handed at age 11 are classified as right handed at age 70. Among those classified as left-handed at age 11; 68·9% were classified as left-handed, 17·8% as mixed handed, and 13·3% as right-handed at age 70. The strong concordance between these results in those assessed as right-handed at age 11 suggests that societal pressure for left-handers to use their right hand during childhood was not a significant factor here. The broader range of results among those classified as left-handed was anticipated given the known spectrum of left-handedness. Among those classified as either left-handed *or* ambidextrous at age 11, there was 85·1% concordance with classification as left-handed or mixed at age 70, supporting our grouping together of those who are left-handed or ambidextrous in the multivariate analyses. It is possible that other factors, including physical impairments may impact hand preference at age 70, while also noting that some aspects of the handedness assessment, such as use of scissors, have a right hand bias.^11^

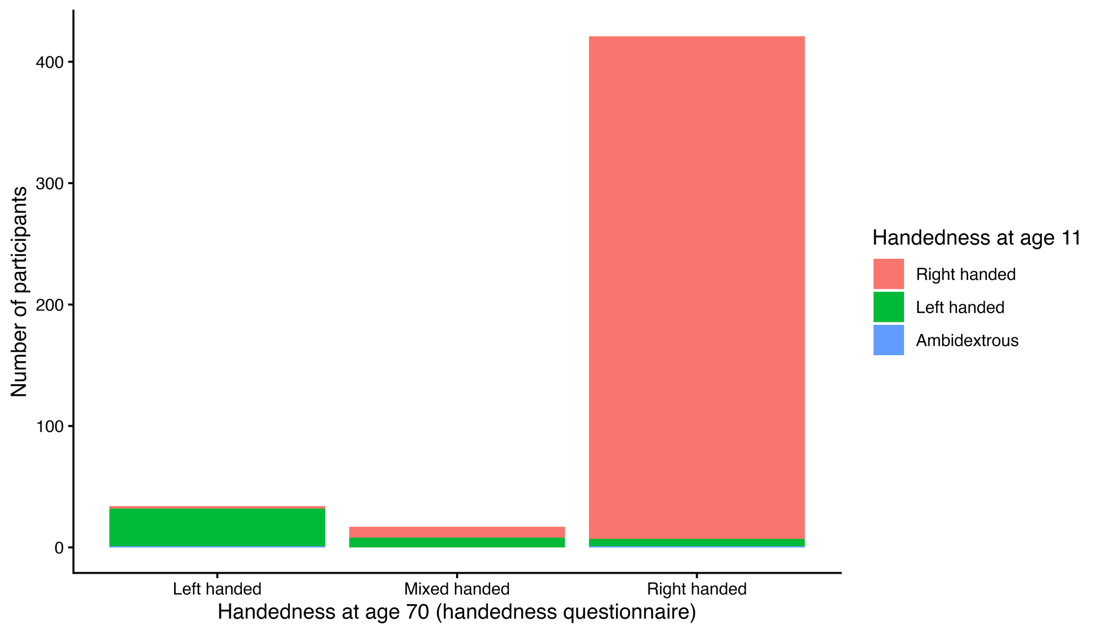

#### Analysis of continuous predictors

Sensitivity analyses were performed for significant results for continuous predictors represented as z-scores to examine whether the linearity assumption was appropriate. z-scores were converted into a three-level factor variable: low (>1 SD below the mean), normal (within ±1 SD of the mean), and high (>1 SD above the mean). Risk was lowest in the low group (>1 SD below the mean), intermediate in the normal group (within ±1 SD of the mean), and highest in the high group (>1 SD above the mean). This suggests associations are not driven solely by individuals with high BMI or severe mental health problems; those with lower levels have correspondingly lower risk. The relationship between emotional problems (z-score) and mortality in males was not monotonic; mortality risk was elevated only for those with a high burden of emotional problems. Hazard ratios (HR) represent the outcome risk for high or low predictor levels relative to normal levels.

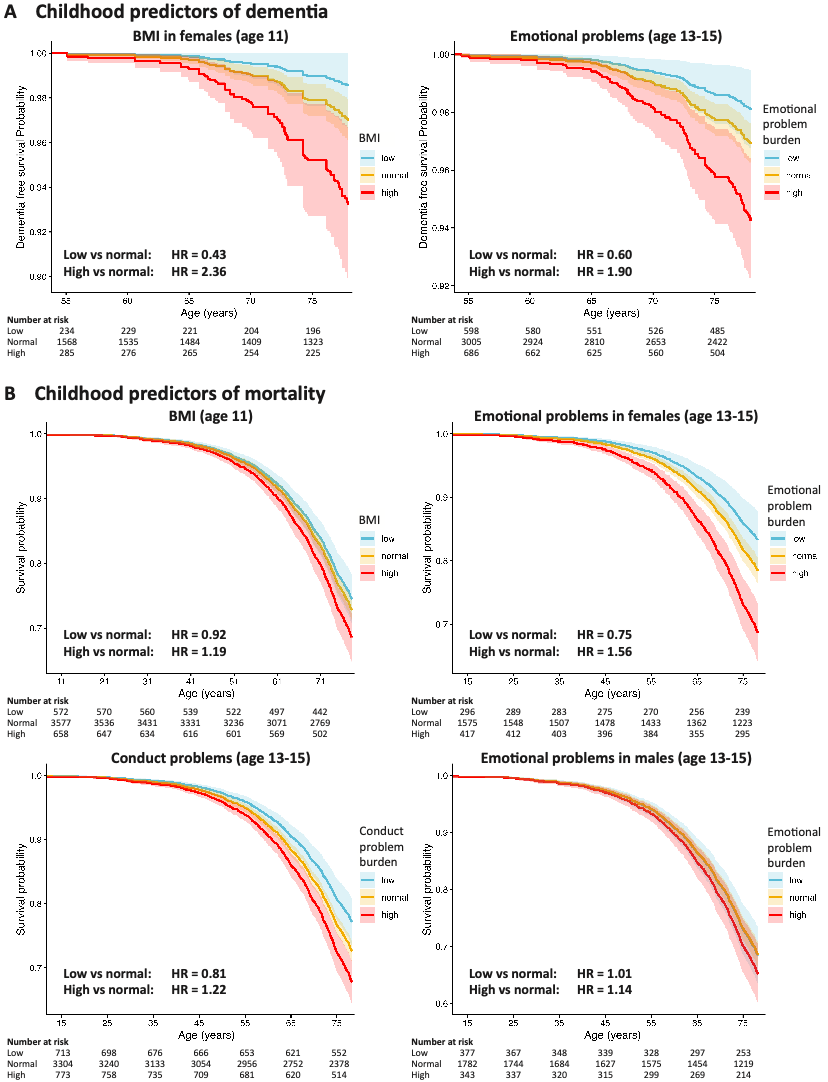

### Appendix 7. Interactions with sex

**Interactions with sex**

Forest plots demonstrating significant interactions with sex in the relationships between *APOE* ε4 carrier status, left-handedness, and BMI and dementia risk (**A**), and mental health (emotional problems), and mortality (**B**). In all cases, the significant relationship between the predictor and dementia/mortality was only present in females.

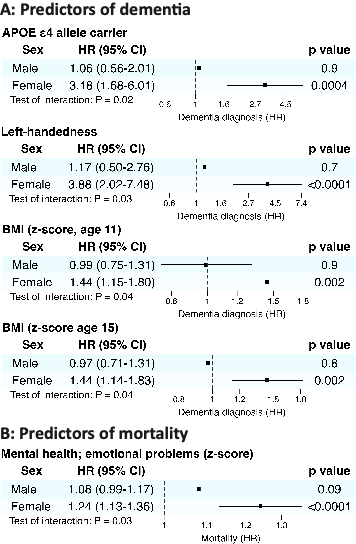

**Sex interactions in relationship between childhood factors and dementia risk.** Table showing strength of the interaction with (female) sex in the relationship between childhood factors, and dementia risk. Reported statistics represent the hazard ratio and 95% confidence interval (CI) for the interaction term only. The interaction with sex in relationship between being ambidextrous or having nocturnal enuresis at age 15 and dementia risk were not calculated (NC)^1^ due to there being too few participants in these subgroups.

| **Predictor** | **Hazard ratio** | **95% CI** | **P value** |
| --- | --- | --- | --- |
| APOE ε4 allele carrier | 3·0 | 1·2 to 7·4 | **0·02** |
| Socioeconomic position | 0·6 | 0·3 to 1·3 | 0·2 |
| Developmental delay |  |  |  |
| ≥ 1 milestone | 2·6 | 0·9 to 7·6 | 0·07 |
| ≥ 2 milestones | 2·0 | 0·5 to 7·8 | 0·3 |
| Cognition |  |  |  |
| Low | 1·0 | 0·4 to 2·5 | 0·9 |
| High | 0·8 | 0·2 to 3·4 | 0·7 |
| Education |  |  |  |
| Any | 0·6 | 0·3 to 1·3 | 0·2 |
| Beyond age 16 | 1·7 | 0·6 to 4·8 | 0·3 |
| Visual impairment | 2·9 | 0·6 to 15·3 | 0·2 |
| Hearing Impairment | 1·8 | 0·3 to 11·2 | 0·5 |
| Pollution exposure | 0·4 | 0·1 to 1·3 | 0·2 |
| Handedness |  |  |  |
| Left-handed | 3·3 | 1·1 to 9·7 | **0·03** |
| Ambidextrous | NC^1^ | | |
| Nocturnal enuresis |  |  |  |
| Age 4 | 0·7 | 0·4 to 1·3 | 0·3 |
| Age 6 | 1·0 | 0·6 to 1·8 | 0·9 |
| Age 8 | 0·7 | 0·3 to 1·7 | 0·4 |
| Age 11 | 0·8 | 0·3 to 2·2 | 0·7 |
| Age 15 | NC^1^ | | |
| Sleep disturbance |  |  |  |
| Age 6 | 0·5 | 0·2 to 1·1 | 0·09 |
| Age 11 | 0·7 | 0·2 to 2·2 | 0·6 |
| Age 16 | 0·5 | 0·2 to 1·5 | 0·2 |
| Mental health (z-score) |  |  |  |
| Emotional problems | 1·2 | 0·8 to 1·8 | 0·4 |
| Conduct problems | 0·9 | 0·6 to 1·3 | 0·5 |
| Birthweight (z-score) | 0·9 | 0·6 to 1·3 | 0·6 |
| BMI (z-score) |  |  |  |
| Age 2 | 1·1 | 0·7 to 1·7 | 0·7 |
| Age 4 | 1·3 | 0·9 to 2·0 | 0·1 |
| Age 6 | 1·1 | 0·7 to 1·7 | 0·6 |
| Age 11 | 1·4 | 1·0 to 2·0 | **0·049** |
| Age 15 | 1·5 | 1·0 to 2·2 | **0·04** |
| Breastfeeding (per 3 months) | 1·0 | 0·7 to 1·3 | 0·8 |

**Sex interactions in relationship between childhood factors and mortality.** Table showing strength of the interaction with (female) sex in the relationship between childhood factors and mortality. Reported statistics represent the hazard ratio and 95% confidence interval (CI) for the interaction term only.

| **Predictor** | **Hazard ratio** | **95% CI** | **P value** |
| --- | --- | --- | --- |
| APOE ε4 allele carrier | 0·8 | 0·6 to 1·2 | 0·3 |
| Socioeconomic position | 1·3 | 1·0 to 1·6 | 0·08 |
| Developmental delay |  |  |  |
| ≥ 1 milestone | 1·3 | 0·9 to 1·9 | 0·1 |
| ≥ 2 milestones | 1·3 | 0·7 to 2·2 | 0·4 |
| Cognition |  |  |  |
| Low | 1·2 | 0·8 to 1·6 | 0·3 |
| High | 0·7 | 0·4 to 1·1 | 0·1 |
| Education |  |  |  |
| Any | 0·9 | 0·7 to 1·1 | 0·4 |
| Beyond age 16 | 1·1 | 0·8 to 1·5 | 0·7 |
| Visual impairment | 0·9 | 0·5 to 1·4 | 0·6 |
| Hearing Impairment | 1·2 | 0·7 to 2·2 | 0·6 |
| Pollution exposure | 0·8 | 0·6 to 1·2 | 0·3 |
| Handedness |  |  |  |
| Left-handed | 0·9 | 0·6 to 1·5 | 0·8 |
| Ambidextrous | 1·7 | 0·4 to 7·7 | 0·5 |
| Nocturnal enuresis |  |  |  |
| Age 4 | 0·9 | 0·7 to 1·1 | 0·3 |
| Age 6 | 1·2 | 1·0 to 1·5 | 0·1 |
| Age 8 | 1·1 | 0·8 to 1·5 | 0·7 |
| Age 11 | 1·3 | 0·9 to 1·8 | 0·2 |
| Age 15 | 0·9 | 0·5 to 1·9 | 0·9 |
| Sleep disturbance |  |  |  |
| Age 6 | 0·9 | 0·7 to 1·2 | 0·6 |
| Age 11 | 1·1 | 0·8 to 1·7 | 0·5 |
| Age 16 | 1·1 | 0·8 to 1·6 | 0·5 |
| Mental health (z-score) |  |  |  |
| Emotional problems | 1·1 | 1·0 to 1·3 | **0·03** |
| Conduct problems | 1.0 | 0·9 to 1·1 | 0·9 |
| Birthweight (z-score) | 1.0 | 0·9 to 1·2 | 0·5 |
| BMI (z-score) |  |  |  |
| Age 2 | 0·9 | 0·8 to 1·1 | 0·3 |
| Age 4 | 0·9 | 0·8 to 1·0 | 0·2 |
| Age 6 | 0·9 | 0·8 to 1·1 | 0·3 |
| Age 11 | 1·1 | 0·9 to 1·2 | 0·4 |
| Age 15 | 1·1 | 1·0 to 1·2 | 0·2 |
| Breastfeeding (per 3 months) | 0·9 | 0·9 to 1·0 | 0·2 |

### Appendix 8: Population attributable fraction.

The population attributable fraction (PAF) estimates the risk of dementia risk and mortality by age 78 under the counterfactual scenario where identified risk factors are not present. The PAF due to individual risk factors, and the total PAF attributable to all risk factors combined, is shown, as well as a total PAF excluding clearly non-modifiable factors (Apolipoprotein E (*APOE*) and sex). BMI; body mass index.

| **Population attributable fraction for dementia risk by age 78** | |
| --- | --- |
| **Exposure** | **PAF** |
| Pollution exposure | 16·5% |
| Mental health: emotional problems | 22·7% |
| *APOE* ε4 carrier status | 21·5% |
| BMI | 14·0% |
| Low cognition | 13·4% |
| Handedness | 9·2% |
| Sleep disturbance | 8·0% |
| Nocturnal enuresis | 5·6% |
| **Total PAF** (all risk factors combined) | **69·5%** |
| Pooled observed risk | 3·42% |
| Pooled expected risk (where all risk factors are not present) | 1·04% |
| **Total PAF** (excluding *APOE* ε4 carrier status) | **61·9%** |
| Pooled expected risk | 1·30% |
| **Population attributable fraction for mortality risk by age 78** | |
| **Exposure** | **PAF** |
| Sex | 17·4% |
| Social class | 11·1% |
| Mental health: emotional problems | 8·3% |
| Education | 10·0% |
| Mental health: conduct problems | 7·4% |
| Breastfeeding | 8·7% |
| BMI | 3·8% |
| Sleep disturbance | 2·3% |
| Nocturnal enuresis | 1·1% |
| **Total PAF** (all risk factors combined) | **56·7%** |
| Pooled observed risk | 27·2% |
| Pooled expected risk (where all risk factors are not present) | 11·8% |
| **Total PAF** (excluding sex) | **43·5%** |
| Pooled expected risk | 15·4% |

**References**

1.	Lane CA, Parker TD, Cash DM, et al. Study protocol: Insight 46 - a neuroscience sub-study of the MRC National Survey of Health and Development. *BMC Neurol* 2017; **17**(1): 75.

2.	Rawle MJ, Davis D, Bendayan R, Wong A, Kuh D, Richards M. Apolipoprotein-E (Apoe) epsilon4 and cognitive decline over the adult life course. *Transl Psychiatry* 2018; **8**(1): 18.

3.	Lu K, Nicholas JM, Collins JD, et al. Cognition at age 70: Life course predictors and associations with brain pathologies. *Neurology* 2019; **93**(23): e2144-e56.

4.	Office of Population Censuses and Surveys. Classification of occupations. *London, UK: HMSO* 1970.

5.	Viner RM, Cole TJ. Who changes body mass between adolescence and adulthood? Factors predicting change in BMI between 16 year and 30 years in the 1970 British Birth Cohort. *Int J Obes (Lond)* 2006; **30**(9): 1368-74.

6.	Richards M, Parsonage M. Childhood mental health and life chances in post-war Britain: insights from three national birth cohort studies (executive summary); 2009.

7.	Pless IB, Douglas JW. Chronic illness in childhood. I. Epidemiological and clinicl characteristics. *Pediatrics* 1971; **47**(2): 405-14.

8.	Colman I, Murray J, Abbott RA, et al. Outcomes of conduct problems in adolescence: 40 year follow-up of national cohort. *BMJ* 2009; **338**: a2981.

9.	Burton MJ, Ramke J, Marques AP, et al. The Lancet Global Health Commission on Global Eye Health: vision beyond 2020. *Lancet Glob Health* 2021; **9**(4): e489-e551.

10.	Douglas JWB, Waller RE. Air Pollution and Respiratory Infection in Children. *British Journal of Preventive & Social Medicine* 1966; **20**(1): 1-&.

11.	Briggs GG, Nebes RD. Patterns of Hand Preference in a Student Population. *Cortex* 1975; **11**(3): 230-8.

12.	Cooper R, Hardy R, Kuh D. Timing of menarche, childbearing and hysterectomy risk. *Maturitas* 2008; **61**(4): 317-22.

13.	Leahy TP, Simpson A, Sammon C, Ballard C, Gsteiger S. Estimating the prevalence of diagnosed Alzheimer disease in England across deprivation groups using electronic health records: a clinical practice research datalink study. *BMJ Open* 2023; **13**(10): e075800.

14.	Brunnstrom H, Englund E. Clinicopathological concordance in dementia diagnostics. *Am J Geriatr Psychiatry* 2009; **17**(8): 664-70.

15.	World Health Organization. International statistical classification of diseases and related health problems. 10th revision, 2nd edition. ed. Geneva: World Health Organization; 2004.

16.	R Core Team. R: A Language and Environment for Statistical Computing. Vienna, Austria: R Foundation for Statistical Computing; 2023.

17.	Bartlett JW, Seaman SR, White IR, Carpenter JR, Alzheimer's Disease Neuroimaging I. Multiple imputation of covariates by fully conditional specification: Accommodating the substantive model. *Stat Methods Med Res* 2015; **24**(4): 462-87.

18.	White IR, Royston P. Imputing missing covariate values for the Cox model. *Stat Med* 2009; **28**(15): 1982-98.

19.	Ferguson J, O'Connell M. Estimating and displaying population attributable fractions using the R package: graphPAF. *Eur J Epidemiol* 2024; **39**(7): 715-42.

20.	Denz R, Klaaßen-Mielke R, Timmesfeld N. A comparison of different methods to adjust survival curves for confounders. *Stat Med* 2023; **42**(10): 1461-79.
